# Cumulative Burden of Prediabetes, Subclinical Myocardial Injury, and Myocardial Stress and Risk of Incident Atrial Fibrillation in Adults With Hypertension

**DOI:** 10.64898/2026.09.02.26362108

**Authors:** Moustafa Elnewishy, Asem M. Mohsen, Tarek Zaho, Richard Kazibwe, Takeki Suzuki, M Benjamin Shoemaker, Prashant D. Bhave, Elsayed Z. Soliman

## Abstract

**Background:** Metabolic dysfunction, subclinical myocardial injury, and myocardial stress may contribute to atrial fibrillation (AF), but ther independent and cumulative associations with incident AF are unclear.

**Methods:** We analyzed 7,261 Systolic Blood Pressure Intervention Trial (SPRINT) participants without prevalent AF, who had baseline fasting glucose, high-sensitivity cardiac troponin I (hs-cTnI), and N-terminal pro–B-type natriuretic peptide (NT-proBNP) measurements. By trial design, SPRINT excluded individuals with diabetes, prior stroke or recent symptomatic heart failure or left ventricular ejection fraction <35%. Prediabetes represented metabolic dysfunction, elevated hs-cTnI myocardial injury, and elevated NT-proBNP myocardial stress. Cox models assessed associations of individual domains and the number of abnormal domains (0–3) with incident AF. Secondary analyses examined the 8 mutually exclusive domain combinations.

**Results:** During a median 3.76-year follow-up, 174 participants developed AF. In multivariable adjusted model, prediabetes, elevated hs-cTnI, and elevated NT-proBNP were associated with incident AF (HR, 1.48 [95% CI, 1.07–2.05], 1.84 [95% CI, 1.30–2.60], and 2.35 [95% CI, 1.56–3.55], respectively). AF risk increased progressively with increasing domain burden (P for trend <0.001); each additional abnormal domain was associated with an 82% higher AF risk (HR, 1.82 [95% CI, 1.50–2.22]). Participants with abnormalities in all 3 domains had the highest risk (HR, 6.11 [95% CI, 2.82–13.24]).

**Conclusions:** Prediabetes, subclinical myocardial injury, and myocardial stress were independently associated with incident AF, with progressively greater risk as abnormalities accumulated. These findings support a multidomain framework in which complementary metabolic and cardiac abnormalities collectively characterize susceptibility to AF.

## Introduction

Atrial fibrillation (AF) is the most common sustained cardiac arrhythmia and results from the convergence of multiple pathophysiologic processes that promote atrial structural and electrical remodeling.^1–3^ Metabolic abnormalities, myocardial injury, hemodynamic stress, inflammation, and fibrosis may contribute to the development of an arrhythmogenic atrial substrate, often before AF becomes clinically apparent.^3^ Multiple cardiovascular risk factors, including obesity, diabetes mellitus, hypertension, and heart failure, are associated with these processes and with the development and progression of AF.^2,3^ However, considerable heterogeneity in AF risk remains among individuals with similar clinical risk profiles, suggesting that assessment of the underlying biological processes may provide additional insight into susceptibility to AF.

Prediabetes represents an intermediate state of dysglycemia that precedes overt diabetes mellitus and may contribute to adverse cardiovascular remodeling.^4^ However, dysglycemia represents only one of several biological processes potentially involved in AF development. Subclinical myocardial injury and myocardial stress may coexist with metabolic abnormalities and reflect complementary aspects of underlying cardiovascular remodeling. These processes can be assessed using circulating cardiac biomarkers: high-sensitivity cardiac troponin I (hs-cTnI) reflects subclinical myocardial injury, whereas N-terminal pro–B-type natriuretic peptide (NT-proBNP) reflects myocardial wall stress and hemodynamic burden.^5,6^ Thus, prediabetes, elevated hs-cTnI, and elevated NT-proBNP may characterize distinct but complementary biological domains relevant to the development of AF.

Prediabetes, elevated cardiac troponin, and elevated natriuretic peptide concentrations have each been associated with incident AF.^7–9^ However, these abnormalities have largely been considered individually, and their relationships with AF when evaluated simultaneously remain less well characterized. Whether metabolic dysfunction, subclinical myocardial injury, and myocardial stress provide independent prognostic information—and, importantly, whether accumulation of abnormalities across these domains is associated with progressively greater AF risk—remains uncertain. Individuals with abnormalities across multiple domains may represent a multidomain high-risk phenotype that is not apparent from any single marker. Defining this phenotype could provide a simple framework for characterizing the biological burden associated with future AF.

Using data from the Systolic Blood Pressure Intervention Trial (SPRINT), we examined the independent and joint associations of prediabetes, elevated hs-cTnI, and elevated NT-proBNP with incident AF. We also examined whether AF risk increased with greater biological-domain burden.

## Methods

### Study Design and Population

This study was a secondary analysis of participants enrolled in the Systolic Blood Pressure Intervention Trial (SPRINT), a multicenter randomized clinical trial comparing intensive versus standard systolic blood pressure treatment. SPRINT enrolled adults aged 50 years or older with hypertension and increased cardiovascular risk but without diabetes mellitus or a history of stroke.^10,11^ SPRINT also excluded participants with symptomatic heart failure within the past 6 months or left ventricular ejection fraction <35% at the time of recruitment.

For the present analysis of incident atrial fibrillation (AF), we excluded participants with prevalent AF on the baseline electrocardiogram (ECG), those without an available follow-up ECG for ascertainment of incident AF, and those with missing baseline fasting glucose measurements because prediabetes status could not be determined. The primary analytic cohort was further restricted to participants with available baseline high-sensitivity cardiac troponin I (hs-cTnI) and N-terminal pro–B-type natriuretic peptide (NT-proBNP) measurements, permitting classification of all 3 biological domains. Participants with missing covariates required for multivariable adjustment were excluded from the corresponding adjusted analyses.

The institutional review boards at all participating centers approved the original SPRINT protocol, and all participants provided written informed consent.

### Assessment of Prediabetes, Subclinical Myocardial Injury, and Myocardial Stress

Three complementary biological domains were evaluated: prediabetes, representing metabolic dysfunction; elevated hs-cTnI, representing subclinical myocardial injury; and elevated NT-proBNP, representing myocardial stress.

Prediabetes was defined according to American Diabetes Association fasting plasma glucose criteria as a fasting glucose concentration of 100 to 125 mg/dL.^12^ Because diabetes mellitus was an exclusion criterion for SPRINT, all participants were free of known diabetes at trial enrollment. Participants with fasting glucose concentrations below 100 mg/dL were classified as not having prediabetes.

Baseline hs-cTnI and NT-proBNP concentrations were measured at Baylor College of Medicine using chemiluminescent immunoassays on the Architect i2000SR automated analyzer (Abbott). hs-cTnI was measured using the Architect Stat High Sensitive Troponin-I assay and NT-proBNP using the Alere NT-proBNP for Architect assay. Subclinical myocardial injury was defined as an hs-cTnI concentration of ≥6 ng/L in men or ≥4 ng/L in women, and myocardial stress as an NT-proBNP concentration of ≥125 pg/mL.^13^

To evaluate the cumulative burden of abnormalities across the 3 biological domains, we calculated a biological-domain burden by assigning 1 point for the presence of each abnormal domain: prediabetes, elevated hs-cTnI, and elevated NT-proBNP. Participants were categorized as having 0, 1, 2, or 3 abnormal domains. This construct was intended to characterize cumulative biological-domain burden rather than serve as a clinical risk score.

For secondary analyses, participants were classified into 8 mutually exclusive groups representing all possible combinations of the 3 domains: no abnormal domains; prediabetes alone; elevated hs-cTnI alone; elevated NT-proBNP alone; prediabetes and elevated hs-cTnI; prediabetes and elevated NT-proBNP; elevated hs-cTnI and NT-proBNP; and abnormalities in all 3 domains. Participants without any of the 3 abnormalities served as the reference group.

### Ascertainment of Incident Atrial Fibrillation

Incident AF was ascertained from protocol 12-lead ECGs obtained at baseline, year 2, year 4, and the closeout visit. ECGs were digitally recorded using GE MAC 1200 electrocardiographs (GE Healthcare) at standard calibration (10 mm/mV and 25 mm/s) and transmitted to the Epidemiological Cardiology Research Center (EPICARE) at Wake Forest University School of Medicine.

ECG quality was reviewed by trained technicians blinded to randomized treatment assignment before automated processing using the GE Marquette 12-SL algorithm (version 2001). ECG abnormalities were classified according to the Minnesota Code.^14^ ECGs classified as AF were visually confirmed by trained ECG readers according to previously described SPRINT procedures.^15^ Participants with baseline AF were excluded from the present analysis, and incident AF was defined by the first follow-up protocol ECG demonstrating AF.

### Covariates

Covariates were selected a priori based on established associations with incident AF and their potential to confound the associations of interest. These included age, sex, race, randomized blood pressure treatment assignment, educational attainment, smoking status, alcohol consumption, physical activity, body mass index, systolic blood pressure, number of antihypertensive medications, total cholesterol to high-density lipoprotein cholesterol ratio, triglyceride level, estimated glomerular filtration rate, urine albumin-to-creatinine ratio, and prevalent cardiovascular disease. Urine albumin-to-creatinine ratio was natural log-transformed because of its right-skewed distribution.

### Statistical Analysis

Baseline characteristics were summarized according to biological-domain burden (0–3 abnormal domains). Continuous variables are presented as mean (SD) or median (IQR), as appropriate, and categorical variables as number (percentage). Differences across groups were evaluated using analysis of variance or the Kruskal-Wallis test for continuous variables and the χ² test for categorical variables, as appropriate.

Cox proportional hazards regression was used to estimate hazard ratios (HRs) and 95% CIs for incident AF. The associations of prediabetes, elevated hs-cTnI, and elevated NT-proBNP with incident AF were first evaluated individually using separate Cox models. Three sequential models were constructed. Model 1 adjusted for age, sex, race, and randomized blood pressure treatment assignment. Model 2 additionally adjusted for educational attainment, smoking status, alcohol consumption, physical activity, body mass index, systolic blood pressure, number of antihypertensive medications, total cholesterol to high-density lipoprotein cholesterol ratio, triglyceride level, estimated glomerular filtration rate, log-transformed urine albumin-to-creatinine ratio, and prevalent cardiovascular disease. Model 3 simultaneously included prediabetes, elevated hs-cTnI, and elevated NT-proBNP together with all Model 2 covariates to evaluate whether each domain remained associated with incident AF after accounting for the other 2 domains.

The association between cumulative biological-domain burden and incident AF was evaluated using fully adjusted Cox models, with participants having no abnormal domains serving as the reference group. Biological-domain burden was also modeled as an ordinal variable ranging from 0 to 3 to estimate the HR associated with each additional abnormal domain and to test for a linear trend across increasing domain burden. To examine whether specific combinations of abnormalities were associated with AF, participants were additionally classified into 8 mutually exclusive groups representing all possible combinations of prediabetes, elevated hs-cTnI, and elevated NT-proBNP. Fully adjusted HRs and 95% CIs were estimated with participants free of all 3 abnormalities serving as the reference group. Because of the smaller number of AF events within some individual combinations, these analyses were considered secondary.

Kaplan-Meier methods were used to estimate the cumulative probability of incident AF according to biological-domain burden and the 8 mutually exclusive biological-domain combinations. Differences among groups were evaluated using the log-rank test, with numbers at risk displayed below each figure.

Because sequential models differed in sample size owing to missing covariate data, a same-sample sensitivity analysis was performed to determine whether changes in the association between prediabetes and incident AF after biomarker adjustment reflected differences in the analytic sample. The fully adjusted clinical model for prediabetes was repeated among the identical complete-case cohort included in Model 3, and the HR for prediabetes was compared before and after additional adjustment for hs-cTnI and NT-proBNP.

All statistical tests were 2-sided, with P<.05 considered statistically significant. Statistical analyses were performed using SAS, version 9.4 (SAS Institute Inc).

## Results

The primary analytic cohort included 7,261 SPRINT participants without prevalent atrial fibrillation (AF) and with complete baseline information on prediabetes, high-sensitivity cardiac troponin I (hs-cTnI), and N-terminal pro–B-type natriuretic peptide (NT-proBNP) (**Figure 1**).

**Figure 1.**
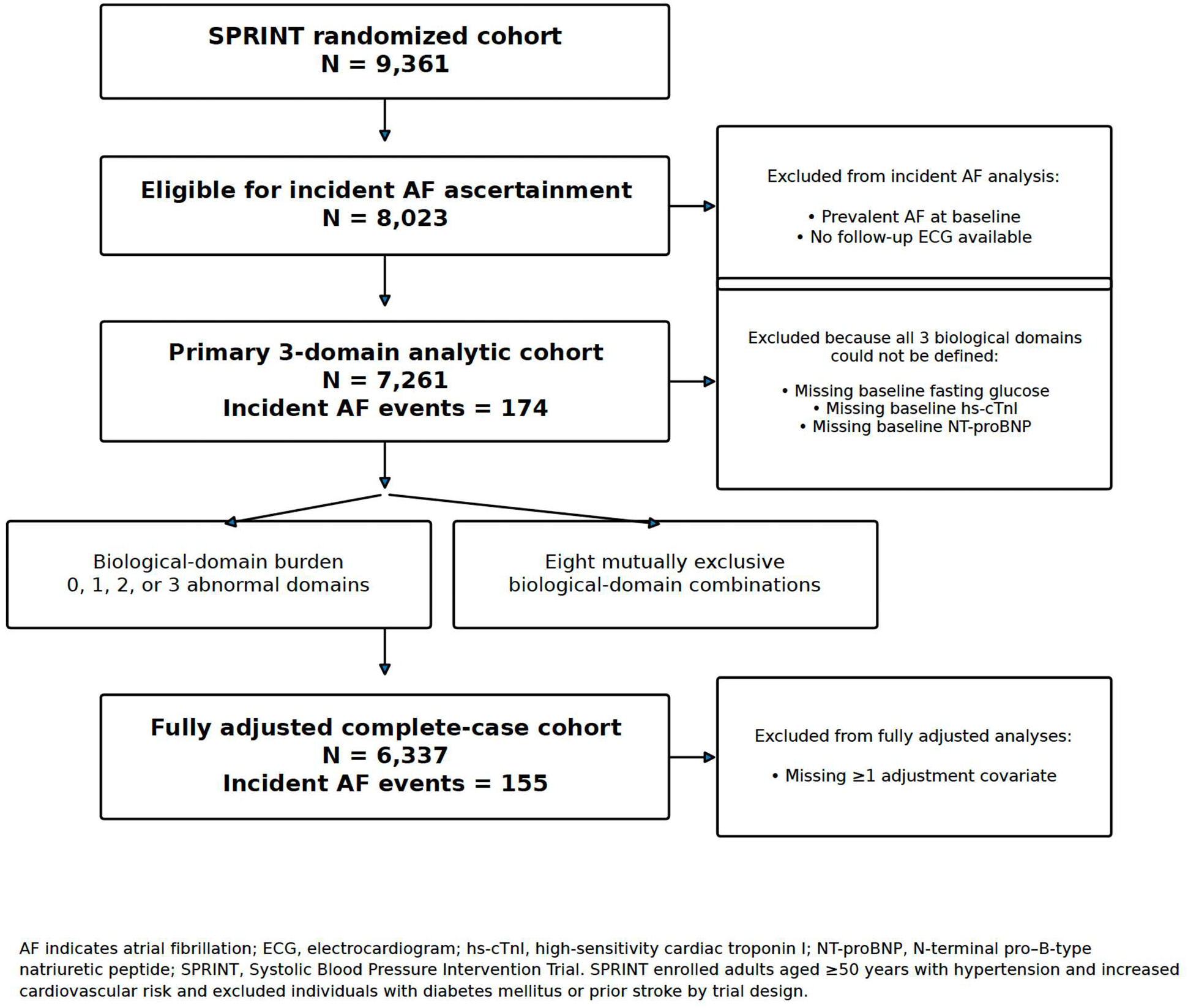
Derivation of the Analytic Cohort

Baseline characteristics according to biological-domain burden are presented in **Table 1**. Participants with a greater number of abnormal biological domains were older and generally had a less favorable cardiovascular risk profile.

**Table 1.** Baseline Characteristics According Biological Domain Burden.

| Characteristic | 0 Domains<br>(n=1803) | 1 Domain<br>(n=2916) | 2 Domains<br>(n=1949) | 3 Domains<br>(n=593) | p-value |
| --- | --- | --- | --- | --- | --- |
| Age (year) | 64.2±8.0 | 67.3±8.8 | 70.5±9.5 | 72.4±9.5 | <0.001 |
| Women | 613 (34.0) | 1033 (35.4) | 790 (40.5) | 225 (37.9) | <0.001 |
| Non-Hispanic Black | 649 (36.0) | 865 (29.7) | 577 (29.6) | 134 (22.6) | <0.001 |
| College education | 755 (41.9) | 1249 (42.8) | 813 (41.7) | 226 (38.1) | 0.206 |
| Current smoking | 280 (16.6) | 308 (11.4) | 193 (10.8) | 49 (9.2) | <0.001 |
| Daily alcohol consumption | 63 (3.5) | 115 (3.9) | 83 (4.3) | 18 (3.0) | 0.451 |
| Regular physical activity | 1306 (77.9) | 2048 (75.8) | 1266 (71.1) | 358 (67.2) | <0.001 |
| BMI (kg/m<sup>2</sup>) | 29.5±5.4 | 29.9±5.8 | 29.7±5.9 | 30.2±5.7 | 0.031 |
| Systolic BP (mm Hg) | 136.7±13.6 | 139.3±15.2 | 142.2±16.6 | 143.2±16.8 | <0.001 |
| Diastolic BP (mm Hg) | 80.3±10.4 | 78.8±11.4 | 76.7±12.7 | 74.3±12.9 | <0.001 |
| Antihypertensive medications. | 2.0 (1.0–2.0) | 2.0 (1.0–2.0) | 2.0 (1.0–3.0) | 2.0 (2.0–3.0) | <0.001 |
| Intensive BP treatment | 891 (49.4) | 1429 (49.0) | 1005 (51.6) | 303 (51.1) | 0.309 |
| Prevalent CVD | 210 (11.6) | 450 (15.4) | 531 (27.2) | 199 (33.6) | <0.001 |
| eGFR (mL/min/1.73 m<sup>2</sup>) | 77.4±18.6 | 73.4±20.0 | 67.0±20.1 | 61.8±20.3 | <0.001 |
| Urine ACR (mg/g) | 7.4 (4.9–13.3) | 8.5 (5.4–17.5) | 11.7 (6.6–29.8) | 15.5 (7.9–44.8) | <0.001 |
| HDL cholesterol (mg/dL) | 53.3±14.6 | 52.8±14.4 | 53.8±14.5 | 52.0±14.1 | 0.025 |
| Total cholesterol (mg/dL) | 195.9±40.7 | 191.4±40.4 | 187.5±41.5 | 182.8±40.9 | <0.001 |
| Triglycerides (mg/dL) | 105.0 (77.0–148.0) | 107.0 (78.0–152.0) | 104.0 (75.0–143.0) | 106.0 (78.0–154.0) | 0.022 |
| Fasting glucose (mg/dL) | 91.0±6.2 | 98.4±10.8 | 98.9±10.6 | 107.6±6.4 | <0.001 |
| Prediabetes | 0 (0.0) | 1355 (46.5) | 985 (50.5) | 593 (100.0) | <0.001 |
| hs-cTnI (ng/L) | 2.8 (2.1–3.7) | 3.5 (2.4–5.0) | 6.6 (4.4–10.2) | 8.6 (6.3–13.1) | <0.001 |
| Elevated hs-cTnI | 0 (0.0) | 574 (19.7) | 1403 (72.0) | 593 (100.0) | <0.001 |
| NT-proBNP (pg/mL) | 58.5 (36.7–86.2) | 89.5 (50.4–155.2) | 196.4 (130.5–332.3) | 304.6 (195.2–507.3) | <0.001 |
| Elevated NT-proBNP | 0 (0.0) | 987 (33.8) | 1510 (77.5) | 593 (100.0) | <0.001 |

ACR indicates albumin-to-creatinine ratio; BP, blood pressure; CVD, cardiovascular disease; eGFR, estimated glomerular filtration rate; HDL, high-density lipoprotein; hs-cTnI, high-sensitivity cardiac troponin I; IQR, interquartile range; NT-proBNP, N-terminal pro-B-type natriuretic peptide; and SD, standard deviation.
Abnormal biological domains were defined as metabolic dysfunction (prediabetes), subclinical myocardial injury (elevated hs-cTnI), and myocardial stress (elevated NT-proBNP). Participants were categorized according to the total number of abnormal domains (0–3).
Continuous variables are presented as mean (SD) or median (IQR), as appropriate; categorical variables are presented as No. (%).
Overall P values were calculated using analysis of variance, the Kruskal–Wallis test, or the $\chi^2$ test, as appropriate.
Percentages are calculated among participants with available data for each characteristic; denominators may therefore differ because of missing data.

During a median follow-up of 3.76 years, 174 participants in the primary analytic cohort developed incident AF. Among the 6,337 participants with complete data for multivariable adjustment, 155 incident AF events occurred. Prediabetes, elevated hs-cTnI, and elevated NT-proBNP were each significantly associated with incident AF after adjustment for demographic, lifestyle, and cardiovascular risk factors (**Table 2**). In the mutually adjusted model, all 3 domains remained independently associated with incident AF: prediabetes (HR, 1.48; 95% CI, 1.07– 2.05), elevated hs-cTnI (HR, 1.84; 95% CI, 1.30–2.60), and elevated NT-proBNP (HR, 2.35; 95% CI, 1.56–3.55).

**Table 2:** Associations of Prediabetes, Elevated hs-cTnI, and Elevated NT-proBNP With Incident Atrial Fibrillation.

| Exposure | Hazard Ratio (95% Confidence Interval) |  |  |
| --- | --- | --- | --- |
|  | Model 1 | Model 2 | Model 3 |
| Prediabetes | 1.50 (1.09–2.06) | 1.42 (1.03–1.97) | 1.48 (1.07–2.05) |
| Elevated hs-cTnI | 2.51 (1.80–3.50) | 2.05 (1.45–2.89) | 1.84 (1.30–2.60) |
| Elevated NT-proBNP | 3.00 (2.03–4.44) | 2.53 (1.69–3.80) | 2.35 (1.56–3.55) |

Model 1: Adjusted for age, sex, race, and randomized treatment assignment.
Model 2: Additionally adjusted for education, smoking, alcohol consumption, physical activity, BMI, systolic BP, number of antihypertensive medications, total cholesterol/HDL cholesterol ratio, triglycerides, eGFR, log-transformed urine ACR, and prevalent CVD.
Model 3: Model 2 plus mutual adjustment for prediabetes, elevated hs-cTnI, and elevated NT-proBNP.

AF risk increased progressively with greater biological-domain burden (**Table 3**). Compared with participants with no abnormal domains, the adjusted HRs for incident AF were 1.78 (95% CI, 0.86–3.71) for 1 abnormal domain, 3.94 (95% CI, 1.92–8.08) for 2 abnormal domains, and 6.11 (2.82–13.24) for all 3 abnormal domains. When biological-domain burden was modeled as an ordinal variable, each additional abnormal domain was associated with an 82% higher risk of incident AF (HR, 1.82; 95% CI, 1.50–2.22; P for trend <.001). Kaplan-Meier estimates of the cumulative probability of incident AF demonstrated progressive separation according to increasing biological-domain burden during follow-up (**Figure 2**).

**Figure 2.**
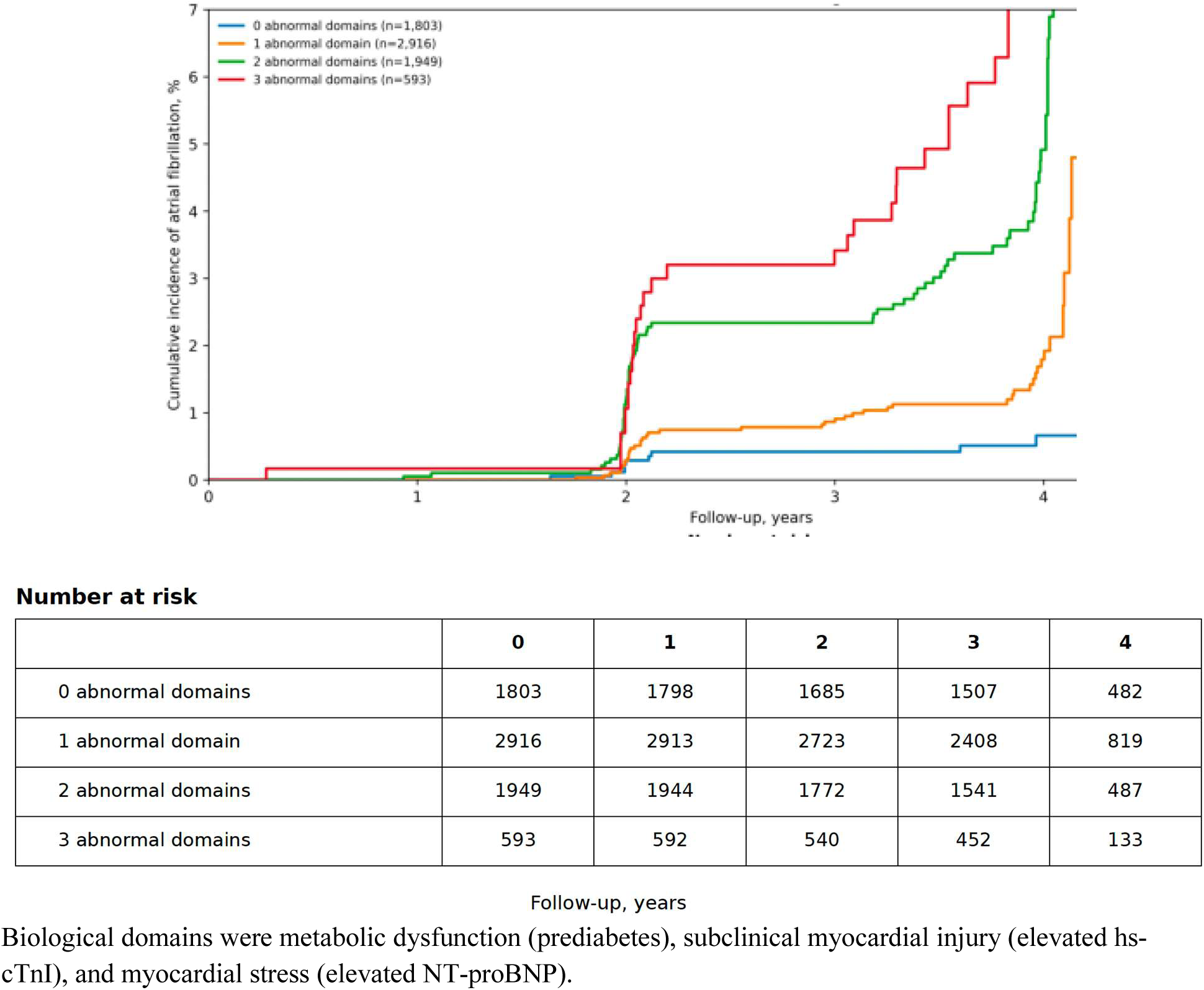
Cumulative Probability of Incident Atrial Fibrillation According to Biological-Domain Burden

**Table 3:** Association of Cumulative Biological-Domain Burden With Incident Atrial Fibrillation.

| Abnormal domains | Participants |  | AF events | Person-years | Rate per 1,000 PY | Adjusted HR (95% CI) | P value |
| --- | --- | --- | --- | --- | --- | --- | --- |
|  | N | % |  |  |  |  |  |
| 0 | 1572 | 24.8 | 9 | 5488.24 | 1.64 | 1.00 (reference) |  |
| 1 | 2554 | 40.3 | 39 | 8937.16 | 4.36 | 1.78 (0.86–3.71) | 0.121 |
| 2 | 1693 | 26.7 | 70 | 5792.14 | 12.09 | 3.94 (1.92–8.08) | <0.001 |
| 3 | 518 | 8.2 | 37 | 1715.32 | 21.57 | 6.11 (2.82–13.24) | <0.001 |

Participants were categorized according to the total number of abnormal domains (0–3). Biological domains were metabolic dysfunction (prediabetes), subclinical myocardial injury (elevated hs-cTnI), and myocardial stress (elevated NT-proBNP)
Model adjusted for: Age, sex, race, and randomized blood-pressure treatment group, college education, smoking, alcohol use, physical activity, BMI, SBP, number of BP medications, total cholesterol/HDL ratio, triglycerides, eGFR, log-transformed UACR, and CVD history.
The fully adjusted analysis included 6,337 participants with complete covariate data and 155 incident AF events
Each additional abnormal domain was associated with an HR of 1.82 (95% CI, 1.50–2.22; P for trend <.001

In secondary analyses, participants were classified into 8 mutually exclusive groups according to the presence or absence of prediabetes, elevated hs-cTnI, and elevated NT-proBNP (**Table 4**). AF risk was generally greater among participants with abnormalities spanning multiple biological domains. Participants with abnormalities in all 3 domains had the highest estimated risk of incident AF compared with those with no abnormalities (adjusted HR, 6.11; 95% CI, 2.82–13.24). Kaplan-Meier estimates similarly demonstrated differences across the 8 biological-domain combinations (log-rank P<.001; **Figure 3**).

**Figure 3.**
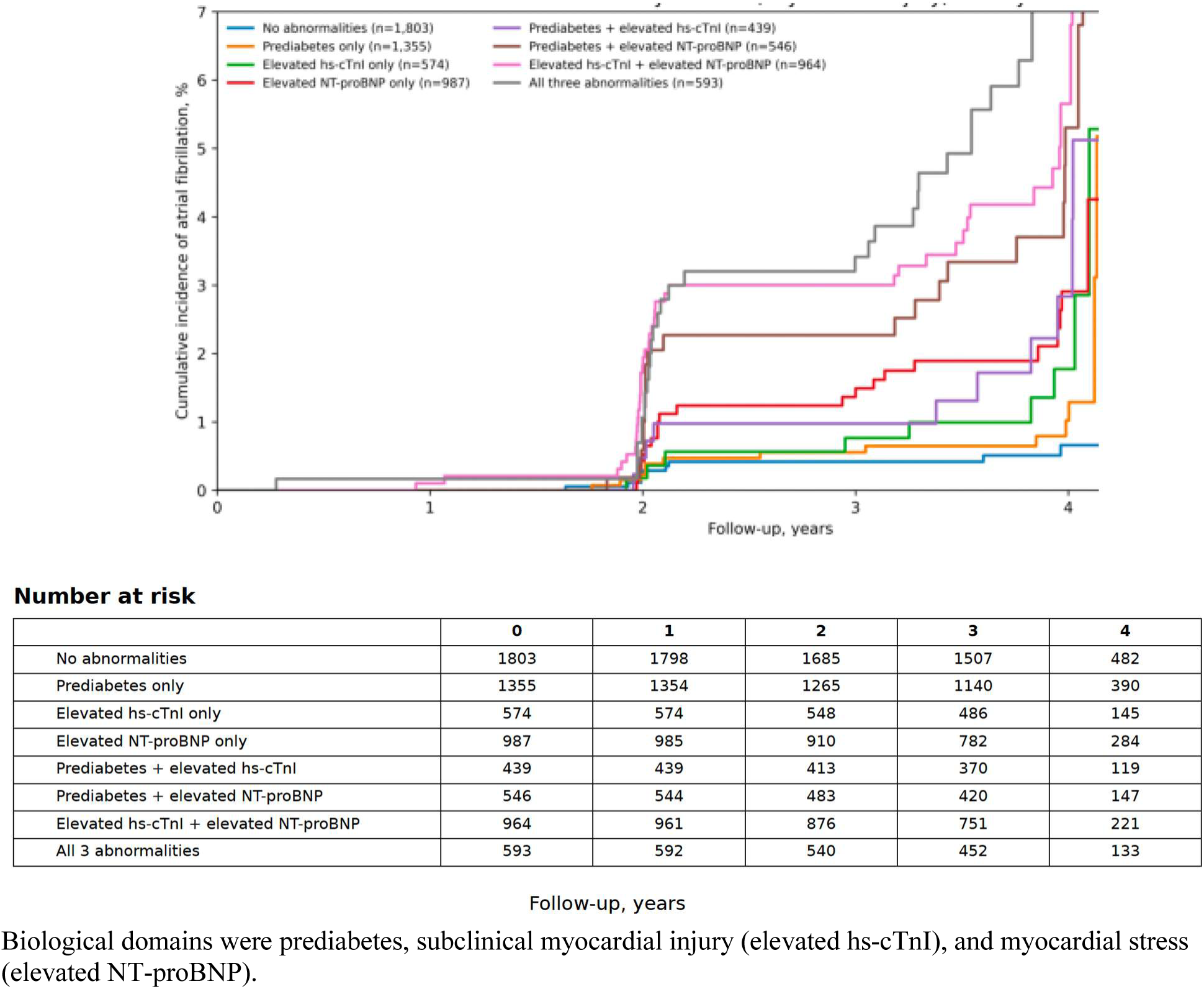
Cumulative Probability of Incident Atrial Fibrillation According to Combinations of Biological Domains.

**Table 4:** Associations of Specific Biological-Domain Combinations With Incident Atrial Fibrillation.

| **Combination** | **N / AF events** | **Adjusted HR (95% CI)** | **P value** |
| --- | --- | --- | --- |
| No abnormalities | 1,572 / 9 | Reference | — |
| Prediabetes only | 1,199 / 12 | 1.35 (0.57–3.20) | 0.502 |
| Elevated hs-cTnI only | 493 / 9 | 2.27 (0.89–5.80) | 0.086 |
| Elevated NT-proBNP only | 862 / 18 | 2.12 (0.94–4.82) | 0.072 |
| Prediabetes + elevated hs-cTnI | 371 / 8 | 2.75 (1.05–7.21) | 0.039 |
| Prediabetes + elevated NT-proBNP | 472 / 21 | 4.33 (1.95–9.63) | <0.001 |
| Elevated hs-cTnI + elevated NT-proBNP | 850 / 41 | 4.47 (2.09–9.56) | <0.001 |
| All three abnormalities | 518 / 37 | 6.11 (2.82–13.24) | <0.001 |

Biological domains were metabolic dysfunction (prediabetes), subclinical myocardial injury (elevated hs-cTnI), and myocardial stress (elevated NT-proBNP)
hs-cTnI, high-sensitivity cardiac troponin I; NT-proBNP, N-terminal pro-B-type natriuretic peptide
Model adjusted for: Age, sex, race, and randomized blood-pressure treatment group, college education, smoking, alcohol use, physical activity, BMI, SBP, number of BP medications, total cholesterol/HDL ratio, triglycerides, eGFR, log-transformed UACR, and CVD history.

In a sensitivity analysis restricted to the same complete-case cohort of 6,337 participants used for the mutually adjusted model, the association between prediabetes and incident AF was similar before and after adjustment for hs-cTnI and NT-proBNP. The adjusted HR for prediabetes was 1.42 (95% CI, 1.03–1.97) before biomarker adjustment and 1.48 (95% CI, 1.07– 2.05) after additional adjustment for both biomarkers.

## Discussion

In this analysis of adults with hypertension, three complementary abnormalities, metabolic dysfunction, subclinical myocardial injury, and myocardial stress, were independently associated with incident AF when evaluated simultaneously. More importantly, AF risk increased markedly with accumulation of abnormalities across these domains, suggesting that multidomain biological burden identifies susceptibility not captured by any single abnormality.

AF is increasingly recognized as the consequence of progressive atrial remodeling arising from multiple interacting biological processes.^16^ Although age, hypertension, obesity, and diabetes are established clinical risk factors for AF, their effects may involve downstream processes including atrial fibrosis, inflammation, oxidative stress, electrical remodeling, and alterations in myocardial structure and function.^17^ Metabolic dysfunction, subclinical myocardial injury, and myocardial stress may therefore reflect complementary components of the biological substrate that precedes clinically apparent AF. The persistence of associations for prediabetes, hs-cTnI, and NT-proBNP after mutual adjustment supports the concept that these domains capture distinct, although potentially overlapping, dimensions of AF susceptibility.

Diabetes mellitus is an established risk factor for AF, but whether earlier stages of dysglycemia are independently associated with AF has been less certain.^9,18^ Previous epidemiologic studies examining prediabetes and incident AF have yielded inconsistent findings, with some associations attenuating after adjustment for conventional cardiovascular risk factors.^19^ In the present study, prediabetes remained independently associated with incident AF after extensive adjustment and after additional adjustment for hs-cTnI and NT-proBNP. In the same complete-case cohort, the association was similar before and after biomarker adjustment, indicating that the persistence of the association was not attributable to differences in the analytic sample. These findings suggest that prediabetes captures a dimension of AF susceptibility not fully reflected by biomarkers of myocardial injury or stress.

The independent associations of hs-cTnI and NT-proBNP further support the multidomain nature of AF risk. Elevated hs-cTnI reflects subclinical cardiomyocyte injury that may accompany hypertensive heart disease, myocardial fibrosis, microvascular ischemia, and other chronic myocardial insults.^20^ In contrast, NT-proBNP reflects myocardial wall stress and hemodynamic burden and may be influenced by increased filling pressures and atrial and ventricular stretch.^21^ Both biomarkers have previously been associated with incident AF in community-based populations.^22,23^ Our findings extend these observations by demonstrating that hs-cTnI and NT-proBNP remained independently associated with AF when evaluated simultaneously with each other and with prediabetes, suggesting that myocardial injury and myocardial stress capture complementary aspects of the biological substrate associated with future AF.

A key finding of this study was the graded association between the number of abnormal biological domains and incident AF. The adjusted risk increased substantially among participants with abnormalities involving 2 or more domains, and participants with abnormalities in all 3 domains had the greatest risk. The 8-group analysis provided further support for this pattern, although estimates for individual combinations were less precise because of smaller numbers of events. These findings suggest that the cumulative burden of abnormalities may be particularly informative: individuals in whom metabolic dysfunction coexists with evidence of subclinical myocardial injury and stress may represent a multidomain high-risk phenotype in whom several processes associated with AF development are already present.

These findings have several potential implications. Simultaneous consideration of metabolic dysfunction, myocardial injury, and myocardial stress may provide a broader characterization of AF susceptibility than consideration of any one domain in isolation. The biological-domain burden used in this study should not, however, be interpreted as a clinical risk score, and we did not test whether it improves discrimination or calibration beyond established AF prediction models. Rather, it provides a simple framework for characterizing the accumulation of abnormalities across complementary biological domains. Individuals with abnormalities spanning multiple domains may represent an enriched population for future studies of intensified risk-factor modification or targeted rhythm surveillance. Whether incorporating these domains into established AF prediction models improves risk stratification and leads to clinically actionable changes warrants further investigation.

This study has several strengths. SPRINT provided a large, well-characterized cohort of adults with hypertension, standardized baseline phenotyping, high-quality biomarker measurements, and protocol-based ECG surveillance with centralized interpretation.

Simultaneous evaluation of prediabetes, hs-cTnI, and NT-proBNP allowed assessment of their independent associations as well as the relationship between cumulative biological-domain burden and AF. Extensive adjustment for demographic characteristics, lifestyle factors, cardiovascular risk factors, kidney function, albuminuria, and prevalent cardiovascular disease, together with the same-sample sensitivity analysis, supported the robustness of the observed associations.

Several limitations should be considered. First, this was a secondary observational analysis of a randomized clinical trial; therefore, the observed associations cannot establish causality, and residual or unmeasured confounding remains possible. Second, the present analysis used baseline hs-cTnI and NT-proBNP measurements and did not evaluate whether longitudinal changes in these biomarkers were associated with AF risk. Third, AF ascertainment was based on intermittent protocol ECGs rather than continuous rhythm monitoring.

Asymptomatic or paroxysmal AF occurring between study ECGs may therefore have gone undetected. Fourth, the number of events was limited in some of the 8 mutually exclusive biological-domain combinations, resulting in imprecise estimates and warranting cautious interpretation of individual combinations. Finally, SPRINT enrolled adults with hypertension at increased cardiovascular risk and excluded individuals with diabetes mellitus and prior stroke by trial design. The findings therefore may not be generalizable to lower-risk populations, individuals with established diabetes, or other populations not represented in SPRINT.

## Conclusions

Among adults with hypertension without diabetes, prediabetes, subclinical myocardial injury, and myocardial stress were independently associated with incident AF. AF risk increased progressively as abnormalities accumulated across these complementary biological domains, identifying a multidomain phenotype at particularly high risk for AF. These findings provide a framework for understanding AF susceptibility as the cumulative burden of metabolic and subclinical cardiac abnormalities and support further investigation of multidomain approaches to AF risk assessment and prevention.

## Data Availability

The SPRINT data used in this analysis are publicly available through the National Heart, Lung, and Blood Institute (NHLBI) Biologic Specimen and Data Repository Information Coordinating Center (BioLINCC) at https://biolincc.nhlbi.nih.

## Acknowledgements

Data for this study were obtained from the publicly available SPRINT data repository through the National Heart, Lung, and Blood Institute (NHLBI) Biologic Specimen and Data Repository Information Coordinating Center (BioLINCC). The present analysis was conducted independently of the SPRINT Research Group, and the conclusions reported herein do not necessarily represent the views of the SPRINT investigators or the NHLBI.

